# The potential impact of declining development assistance for healthcare on population health: projections for Malawi

**DOI:** 10.1101/2024.10.11.24315287

**Authors:** Margherita Molaro, Paul Revill, Martin Chalkley, Sakshi Mohan, Tara Mangal, Tim Colbourn, Joseph H. Collins, Matthew M. Graham, William Graham, Eva Janoušková, Gerald Manthalu, Emmanuel Mnjowe, Watipaso Mulwafu, Rachel Murray-Watson, Pakwanja D. Twea, Andrew N. Phillips, Bingling She, Asif U. Tamuri, Dominic Nkhoma, Joseph Mfutso-Bengo, Timothy B. Hallett

## Abstract

Development assistance for health (DAH) to Malawi will likely decrease as a fraction of GDP in the next few decades. Given the country’s significant reliance on DAH for the delivery of its healthcare services, estimating the impact that this could have on health projections for the country is particularly urgent. We use the Malawi-specific, individual-based “all diseases – whole health-system” *Thanzi La Onse* model to estimate the impact this could have on health system capacities, proxied by the availability of human resources for health, and consequently on population health outcomes. We estimate that the projected changes in DAH could result in a 7-15.8% increase in disability-adjusted life years compared to a scenario where health spending as a percentage of GDP remains unchanged. This could cause a reversal of gains achieved to date in many areas of health, although progress against HIV/AIDS appears to be less vulnerable. The burden due to non-communicable diseases, on the other hand, is found to increase irrespective of yearly growth in health expenditure, if assuming current reach and scope of interventions. Finally, we find that greater health expenditure will improve population health outcomes, but at a diminishing rate.

## 1. Introduction

Malawi has made remarkable progress in health in the last few decades, achieving a 18-year increase in life expectancy at birth in the country between 2000 and 2021 [1]. Many of these gains were achieved with the significant support of international donors, who contributed up to ∼55% of total health expenditures in the country in 2018/2019 [2]. However, the Institute for Health Metrics and Evaluation (IHME) is forecasting a reduction in the contribution of development assistance for health (DAH) as a fraction of the country’s Gross Domestic Product (GDP) in the next few decades, with the rising contribution of government expenditure on health not expected to increase fast enough to compensate for the shortfall [3].

This would happen at a crucial time for Malawi, which has only recently made substantial, yet potentially reversible, gains in major infectious disease pandemics such as HIV, TB, and malaria. In addition, a rise in the contribution of non-communicable diseases (NCDs) to the country’s health burden means the healthcare system is facing a double burden of infectious diseases and NCDs [4, 5].

In this analysis, we translate a number of hypothetical long-term health expenditure scenarios for Malawi into equivalent levels of human resources for health capacity, and make use of the Malawi-specific “all diseases – whole health – system” *Thanzi La Onse* (www.tlomodel.org, *[6]) model to:*

- Estimate the health burden, quantified in disability adjusted life years (DALYs), between 2019 – 2040 under different levels of annual growth rates in health expenditure over the same period, including those forecasted by the IHME;
- Disaggregate these health burden estimates by causes of illness, to understand the divergent trajectory of these causes under different levels of annual growth rates in health expenditure.

## 2. Method

### 2.1. Overview

The individual-based model *Thanzi La Onse* (TLO, www.tlomodel.org, [6]) simulates the evolution of the health burden in Malawi while capturing: its demographic growth; the evolving incidence of all major risk factors, infectious diseases, NCDs, and comorbidities; the health-seeking and treatment-adherence behaviour of those at risk of or affected by a medical condition requiring care; the range of potential interventions available to both prevent and treat those medical conditions, as well as the effectiveness of these interventions; the accuracy of referral and diagnostics; and the extent to which health services are able to meet the demand for care in all its requirements, including timely access to relevant human resources for health (HRH) and consumables. All these factors are modelled explicitly and self-consistently, and are extensively calibrated to available data in the period 2015 – 2019 (see [6] for details).

We use this model to simulate the health burden that would be incurred under different scenarios of health expenditure in Malawi between 2019 and 2040 (inclusive). Each expenditure scenario is assumed to result in an expansion of human resources for health (HRH) in the country from capabilities initially calibrated to 2018, as discussed in detail in section 2.2. Because the ability of the healthcare system to meet the demand for care in the model is constrained by the HRH available, this allows us to estimate the return in health from each expenditure scenario.

Indeed, given realistic representations of i) the patient-facing time available from each medical cadre at each simulated facility, and ii) the time required from different medical cadres by each type of appointment, both adjusted to available data on productivity levels and available resources, the TLO model ensures that, on each day, care can only be dispensed until patient-facing time has been exhausted (see Appendix A). Failure to receive care results in an increased probability of adverse health outcomes for the individual and, in the case of infectious diseases, of further infection spread among the population, accurately capturing repercussions of HRH constraints on the overall health burden.

In this analysis, only the relationship between expenditure in HRH and health outcome is directly captured (see section 4.3), while consumable availability is assumed to be perfect. We perform a sensitivity analysis on the consumable availability assumption in section Appendix B.

### 2.2. Health expenditure scenarios

We assume that the expansion of HRH capabilities in the country matches the combined growth of GDP and fraction of GDP allocated to healthcare expenditures (*f*_HE_) through combined government and DAH efforts. This assumption is reviewed in detail in section 4.

For simplicity and to facilitate interpretability, we assume that annual fractional changes in GDP and *f*_HE_, referred to as *g*_GDP_ and *g*_fHE_ respectively, are constant for the entire simulated period, although fluctuations are expected in practice. HRH capabilities available in any year *i* can therefore be expressed relative to those in the previous year as:

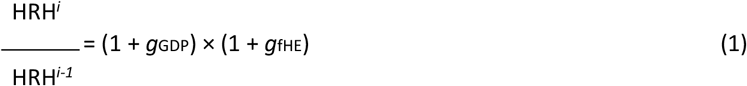

where therefore (1 + *g*_GDP_) × (1 + *g*_fHE_) constitutes the yearly percentage growth of the overall health expenditure, and consequently an equal percentage growth in expenditure on HRH and patient-facing time available, and assume expansion of capabilities starts in the year *i* = 2019 (see Appendix A for details). The expansion of resources due to different health expenditure scenarios can therefore be thought of as an increase in the total amount of medical officers’ patient-facing time available at each facility, resulting in the healthcare system being able to dispense a higher amount of care. Finally, we assume the proportional expansion of available capabilities is the same for all medical officer types, at all facility types, and in all districts. All these assumptions are reviewed in section 4.3.

In this analysis, the HRH expansion scenarios considered are determined by different expectations around how the GDP and *f*_HE_ will grow with time. The range of scenarios considered are summarised and motivated in Table 1. Our worst-case scenario is one where capabilities are not expanded at all from those in 2018 (“No growth” scenario). In all other cases, we assume a fixed GDP growth per year of *g*_GDP_ = 4.2%, which corresponds to the average annual percentage growth of GDP in Malawi (expressed in constant 2015 US$) between 1960 and 2020 according to World Bank data. Two of the scenarios considered (“<< GDP growth” and “< GDP growth”) specifically approximate the IHME forecasts, as discussed in Appendix C.

**Table 1:**
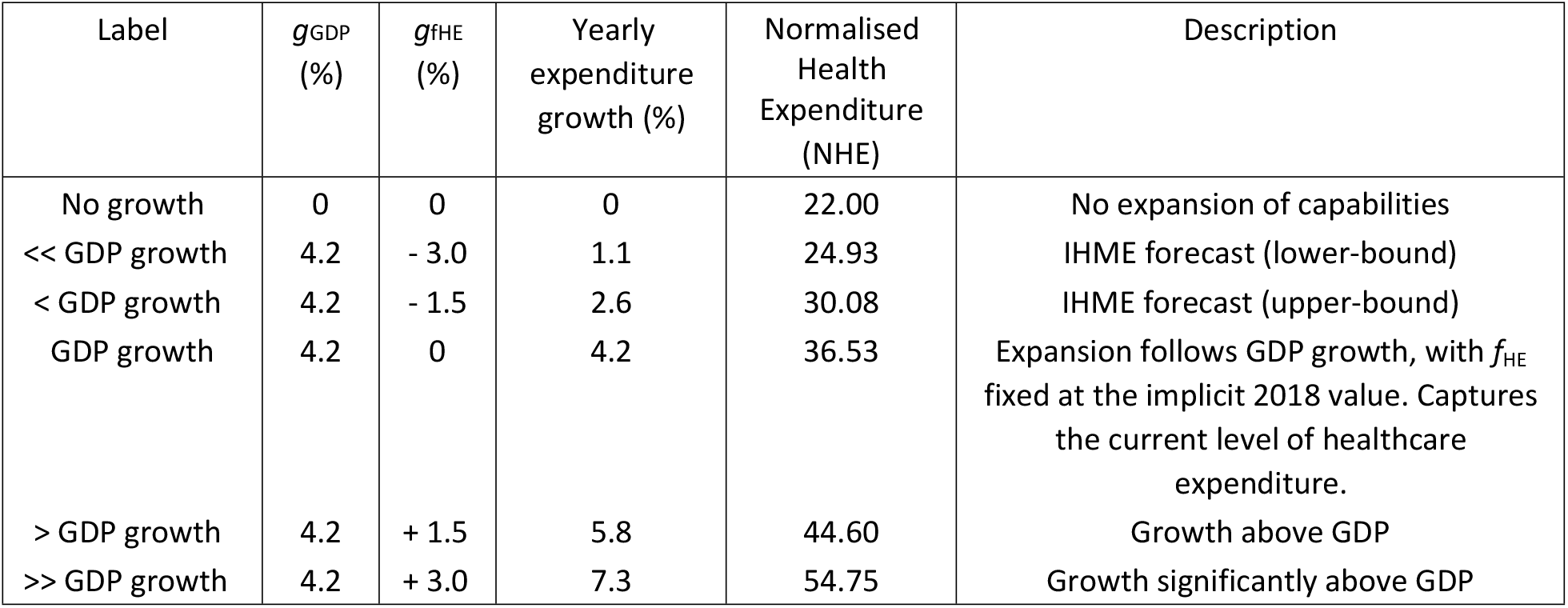
Capabilities expansion scenarios considered. *g*_GDP_ and *g*_fHE_ refer to the growth rate of annual GDP (in constant 2015 US$) and growth rate of fraction of GDP allocated to health (*f*_HE_) assumed, leading to a yearly expenditure growth (%) per scenario of [(1+*g*_GDP_/100)×(1+*g*_fHE_/100)− 1]/100. The normalised health expenditure (NHE) associated with each scenario (defined in Eqn.2) is also included. Scenarios capturing forecasts by the Institute for Health Metrics and Evaluation (IHME) are also included, and discussed in more detail in section Appendix C.

Each scenario *s* is characterised by a different normalised health expenditure (NHE) incurred over the relevant period. We define this in dimensionless units relative to HRH expenses in 2018 as:

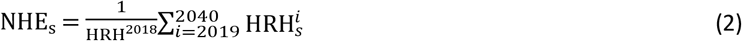

Finally, the health outcome of each health expenditure scenario is quantified by disability-adjusted life years (DALYs) assuming a life-expectancy [7] of 70 years and adopting disability adjustment factors from [8]. The initial population size assumed in 2010 is of 100,000 individuals, while results reported are scaled to the true population size in Malawi in 2010 of 14.5 million individuals. Each scenario considered was simulated 10 independent times, each with independent random draws; both numbers were found to be large enough to give stable estimates of the mean and variance of the health burden obtained under each cause included in the simulation over independent realisations.

### 2.3. Ethics Statement

The *Thanzi La Onse* project received ethical approval from the *College of Medicine Malawi Research Ethics Committee* (COMREC, P.10/19/2820) in Malawi. Only publicly available anonymised secondary data is used in the *Thanzi La Onse* model; therefore, individual informed consent was not required.

## 3. Results

### 3.1. How does health scale with expenditure on human resources for health?

A steady rise in yearly DALYs incurred is observed in the long-term for all scenarios considered (Fig. 1, left plot), suggesting that an expenditure above the largest yearly expenditure growth considered in this analysis (“>> GDP growth”) would be required to stabilise the long-term health burden in the country. This is partly driven by population growth, as illustrated by the equivalent evolution of the life expectancy in the right plot of the same figure: the lowest level of expenditure considered (“No growth” scenario) is indeed still able to stabilise the average life expectancy over the whole period, however failing to achieve any improvements in individual outcomes.

**Figure 1.**
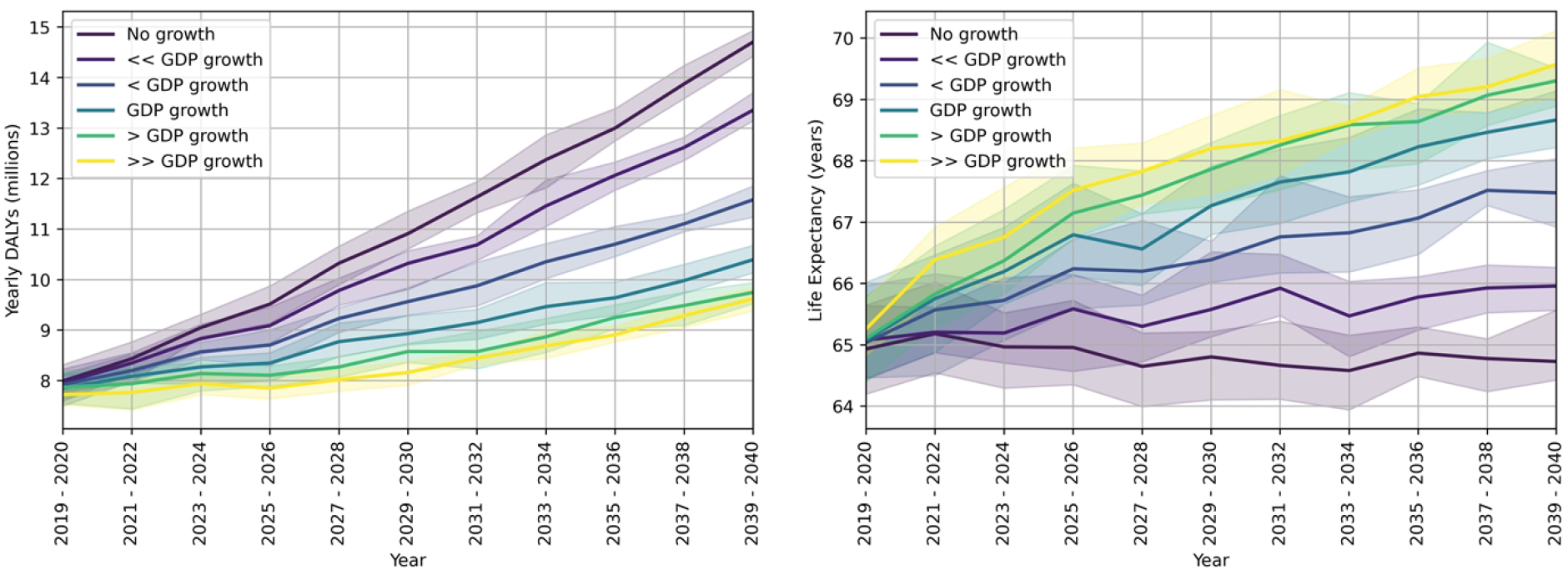
*Left plot*: Total Yearly DALYs (averaged over two-year periods) incurred under different expenditure scenarios. *Right plot*: Life expectancy (averaged over two-year periods) achieved under different expenditure scenarios.

In Fig. 2 (left plot), on the other hand, we show the total DALYs incurred between 2019 and 2040 as a function of the yearly health expenditure growth, as well as the normalised total health expenditure (NHE, defined in Eqn. 2) characterising each expenditure scenario. In the right plot of the same figure, we show the percentage of total DALYs averted compared to the “GDP growth” scenario, which captures the current level of health expenditure.

**Figure 2.**
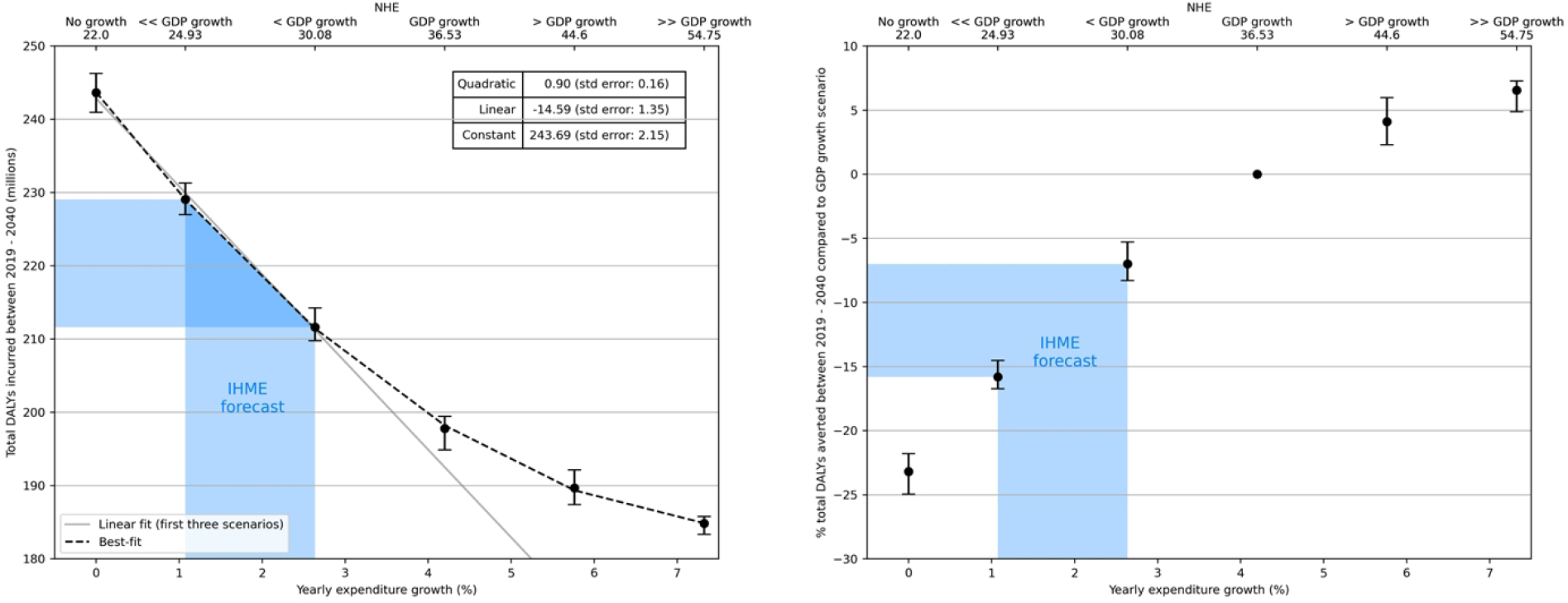
*Left plot:* Total DALYs incurred in the period 2019–2040 (inclusive) as a function of the yearly expenditure growth, as well as the normalised total expenditure (NHE) over that period under each scenario (top x-axis), as defined in Eqn. 2. The best-fit parameters for the function shown are summarised in the table inside the plot, while the linear best-fits to the first three levels of yearly expenditure growth considered are only included for visual guidance. *Right plot:* Percentage DALYs averted in the same period compared to the “GDP growth” scenario, where this captures the current level of health expenditure as a fraction of GDP. The blue shaded area shows the IHME forecast range.

While the reduction in health burden initially achieved is around ∼10 million DALYs for every percentage point increase in expenditure growth, this trend becomes sublinear as the overall expenditure increases above ∼4%, eventually flattening around 8-9%. In particular, a reduction in *f*_HE_ with rates predicted by the IHME (shown by the blue shaded area in the figure) would result in an excess of between 15.8 (14.5 – 16.7 at the 95% confidence interval (CI)) and 7.0 (5.3 – 8.3 at 95% CI) percentage increase, respectively, in total DALYs incurred compared to a fixed *f*_HE_ (i.e. current level of health expenditure) scenario (“GDP growth” scenario).

### 3.2. How are key areas of health affected?

In Fig. 3 we show how three important areas of health are affected by different expenditure strategies, namely:i) major infectious diseases primarily supported via vertical programmes, namely HIV/AIDS, TB, and malaria (HTM); ii) Reproductive, Maternal, Neonatal, and Child Health, including lower respiratory infections, childhood diarrhoea, maternal disorders, measles, and neonatal disorders (RMNCH); and iii) non-communicable diseases, including chronic obstructive pulmonary disease (COPD), cancers, depression/self-harm, diabetes, epilepsy, heart disease, kidney disease, and stroke (NCDs).

**Figure 3.**
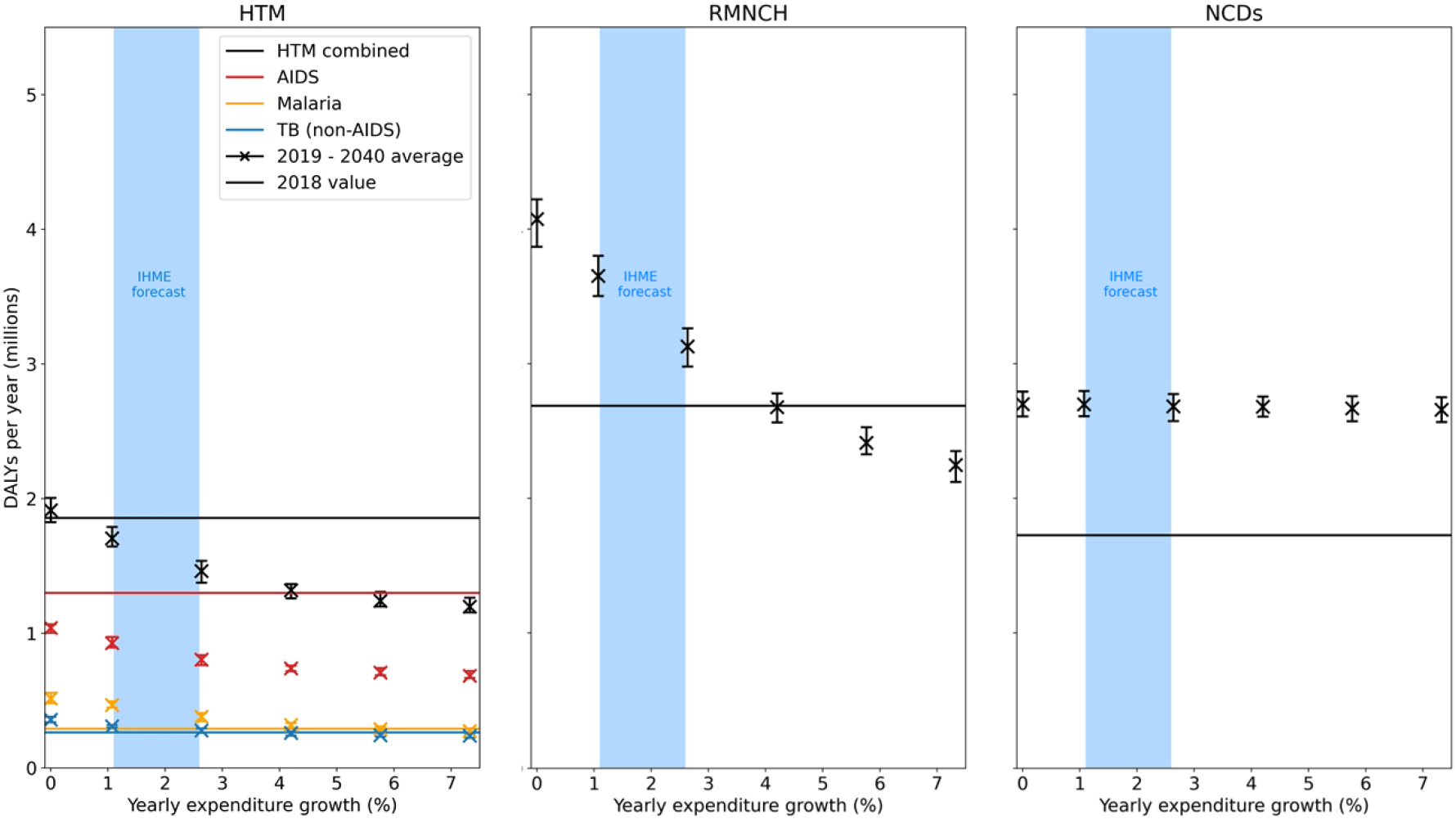
Average yearly DALYs incurred between 2019-2040, grouped into three meaningful categories: HTM, including HIV/AIDS, TB, and malaria; RMNCH, including lower respiratory infections, childhood diarrhoea, maternal and neonatal disorders, and measles; and NCDs, including COPD, cancers, depression/self-harm, diabetes, epilepsy, heart and kidney disease, and stroke. In the case of HTM, DALYs in this area are additionally shown broken down by individual causes. The straight lines indicate the yearly DALYs burden for each cause in 2018. This means that any scenario *above* the respective 2018 level incurred, on average, a worsening of the health burden due to that cause over the 2019-2040 period, whereas any scenario *below* the respective 2018 level incurred an improvement.

Each panel shows the average yearly DALYs lost under each area of health between 2019 – 2040 as a function of the yearly expenditure growth associated with that scenario, while the horizontal lines show the yearly DALYs incurred due to that cause in 2018, the year before scenarios start diverging. This means that if the DALYs incurred under a scenario fall below the respective horizontal line, an average decline in the health burden from that cause was achieved over the period for that scenario. On the other hand, if the DALYs incurred lie above the horizontal lines, the expenditure scenario led to an average increase in the health burden due to that cause between 2019 and 2040.

A downward trend in DALYs incurred due to HIV/AIDS appears to be achievable by all expenditure scenarios, despite significant population growth over this period, suggesting that the important gains made in this area in previous years can be sustained by existing capabilities (although the increase in average yearly DALYs with diminishing yearly expenditure growth suggests that such gains may still be reversed if existing capabilities were to be reduced). The rise in malaria and TB burden would also appear to be mostly contained, but requiring a minimum of “GDP-growth” level expenditure (therefore above IHME projections) to be at least stabilised.

On the other hand, DALYs due to RMNCH would, under IHME projections, significantly rise compared to their 2018 level over the period considered, mainly driven by the population growth over this period. An increase in HRH availability in line with ‘GDP growth” and above appears to be effective at containing this rise and leading, for expenditures above “GDP growth”, to a downward trend in DALYs incurred in this area of health despite an increase in population size.

Finally, and unlike for other areas of health, the rising health burden due to NCDs appears to be largely unaffected by the availability – and therefore expansion – of HRH capabilities. This is due to the complex challenges currently preventing effective NCD programme delivery in the country [9, 10, 11]. Notice that this unconstrained rise in NCDs is therefore what drives the long-term rise in the overall health burden even in the “>> GDP growth” expenditure scenario, which can be observed in Fig. 1.

## 4. Discussion

We have estimated the trends in long-term health-burden that can be expected in Malawi under different scenarios of healthcare expenditure in the period 2019 – 2040, approximated by an equivalent human-resources-for-health (HRH) capabilities expansion. We found that:

- The decline in total DALYs incurred between 2019–2040 is initially close to ∼10 million DALYs per percentage point increase in yearly expenditure growth, but exhibits diminishing returns above 4%. Reasons for these diminishing returns are independent of assumptions around consumable availability (see section Appendix B) and likely linked to: the assumed fixed range of preventative, screening, and treatment serviced offered by the healthcare system over the entire period, which could instead be expanded to include less cost effective options once the most urgent medical needs in the population have been met; persisting difficulties in health service access by certain sections of the population; imperfect diagnostic and referral accuracy; and the intrinsic (less than perfect) effectiveness of each intervention, which are all factors explicitly captured in the TLO model.
- If IHME forecasts of the decline in the fraction of GDP allocated to healthcare from combined governmental and international aid efforts should realise, the country would incur a percentage increase in total DALYs lost of between 15.8 (14.5 – 16.7 at the 95% confidence interval (CI)) and 7.0 (5.3 – 8.3 at 95% CI) percent compared to current levels of health expenditure, under the IHME’s lower and upper bound projections respectively.
- Furthermore, if IHME forecasts of health expenditure were to be realised, the significant gains made by Malawi in the past in important areas of health such RMNCH, malaria, and TB could be reversed. Sustaining or exceeding current levels of health expenditure, on the other hand, would ensure the burden due to these causes can continue to decrease over time despite population growth. In the case of HIV/AIDS, progress made to date – which include the country already meeting two of the three targets set out by USAIDS for 2030 [12] – appears to have made the decline in this burden sustainable by current capabilities.
- The significant rise in the health burden due to NCDs is unaffected by the level of expenditure in HRH, suggesting that expanding the reach and scope of preventative, screening, and treatment services offered would play a key role in ensuring that health investments can effectively translate into a reduction of the health burden due to these causes of ill health in the future [9, 10, 11].

Below, we review some of the assumptions adopted in this analysis.

### 4.1. Assumed incidence of NCDs

A large number of complex factors will shape the incidence of NCDs in the future [4]. The model explicitly accounts for many of these, such as level of wealth, education, sugar, salt, alcohol intake, and body mass index, and does so by extrapolating current trends on the evolution of the incidence of these factors into the future ^1^ [13, 6]. Of course, significant uncertainty remains around such extrapolations, as well as uptake and effectiveness of treatment. By highlighting how significant the future health-burden from such causes would be, should current trends be maintained, this analysis highlights the urgency of implementing preventative as well as curative strategies to ensure both the incidence and the health-cost from these NCDs can be contained in the future.

### 4.2. GDP growth projection

Any projection as far into the future as considered in this analysis is bound to be highly uncertain. Since in our analysis the GDP growth is compounded with *f*_HE_ growth assumptions to describe the overall expansion of capabilities, each scenario can be reinterpreted as representing a different rate of GDP growth by simply rescaling the *g*_fHE_ factor assumed to be describing that scenario, providing some flexibility in the interpretation of the results of these scenarios to include alternative estimates of GDP growth, which however would still be assumed to be constant with time. For example, the scenario “< GDP growth” considered (*g*_GDP_ = 4.2% and *g*_fHE_ = -1.5%) could equivalently describe a *g*_GDP_ = 2% and *g*_fHE_ = 0.62% scenario.

### 4.3. Assumptions around HRH expansion scenarios

In assuming that the capabilities expansion considered in this analysis scaled simply with GDP and *f*_HE_ growth, we implicitly made the following assumptions:

- The fraction of the total health expenditure (GDP × *f*_HE_) allocated to HRH (as opposed to new infrastructure, the purchasing of consumables and equipment, administrative costs, etc.) is constant with time, while the *real costs* of HRH are also assumed to be constant.
- The distribution of HCWs across different cadres, facility levels, and districts remains constant as HRH capabilities are expanded. This strategy for resource allocation may be far from optimal. A more targeted expenditure in particular districts/facility levels/types of cadres may indeed result in a higher return in health from the same expenditure. This analysis should therefore be seen as a “lower bound” to the health benefit that could be obtained from the same expenditure.

In addition, we assumed that any expenditure – based on the above-listed assumptions – directly translated into an expansion of available patient-facing time. In this simplified approach, we therefore did not account for i) possible variations in HCW productivity due to the expansion of existing cadres or other influencing factors, and ii) the fact that the realisation of such scenarios would require adequate and timely expenditures in training and recruitment of additional HCWs, as well as potentially the expansion of existing infrastructures, facilities, and equipment to accommodate the intake of new personnel. These costs may amount to a higher share of the healthcare budget currently allocated to these areas; fully capturing them while imposing a cap on overall expenditure may therefore limit the amount of HRH capabilities expansion achievable under the same assumed GDP and *f*_HE_ growth, as would potential wage increases over the simulated 22-year period. Once such additional costs are captured, such considerations could however easily be embedded in this analysis by refactoring the assumed growth/total expense incurred under each scenario considered. We further do not capture potential variations in health-care workers productivity

Furthermore, while HRH constitutes one of the most important constraints to healthcare delivery and universal healthcare access [14, 15] we do not capture how other constraints -such as consumables, ambulances, and others would scale with increasing expenditure. While in this analysis we assumed perfect consumable availability, results were however found to be largely independent of this assumption (see section Appendix B). In addition, all health services were assumed to be competing equally for the same limited resources. In reality, different funding streams may result in a higher share of overall capabilities being reserved for specific programmes, e.g. through vertical funding ([16]). This share, however, would likely evolve over the period considered as a result of a decline in the contribution of DAH.

In this analysis we were agnostic as to how much of the assumed investment into HRH would be contributed by DAH vs the government of Malawi (GoM). The GoM currently covers the highest proportion of HRH spending (68%) [17]. Health worker salaries and benefits account for 70% of this cost, and are covered at 90% by the GoM. Assuming this present-day allocation of costs, it may be argued that a contraction in international donors support may not affect HRH as significantly as assumed in these scenarios. It is however not unreasonable to assume that the GoM may be forced to reallocate some of its HRH funding were international donors to withdraw their support in other areas of health, resulting in an equivalent contraction. Furthermore, the financial strain placed by a decline in donor support could have a detrimental impact on staffing levels, medical consumable supply, and technical capacity, as experienced in other countries which went through a similar funding transition [18]. Our analysis does not capture these potential effects.

Finally, this analysis does not capture the contribution of prepaid private and out-of-pocket health expenditures to the overall health outcome of the population. The IHME forecasts that the relative contribution of these types of health expenditures to the overall health expenditure in the country will grow over this period from an estimated 17% in 2019 (see Fig. C.6).

## 5. Conclusions

This analysis provided a first-ever quantification of the potential overall consequences of different potential health expenditure scenarios in Malawi in the future, described by an equivalent expansion of human resources for health. It demonstrated the potential risk of reversing gains in several key areas of health in the country if current forecasts on the decline of contribution from development assistance for health were to be realised, and highlighted the need for domestic and international authorities to act in response to this predicted trend.

## Data Availability

The Thanzi La Onse model is open source and available for review and usage at https://github.com/UCL/TLOmodel. In particular, the outputs analysed in this study can be reproduced from model tag "Molaro_et_al_Impact_of_Declining_DAH" (accessible at https://github.com/UCL/TLOmodel/tags).

## Appendix A. Modelling HRH constraints

The TLO model self-consistently captures the number of specific appointment types (referred to as “health-system interactions” or HSIs) requested on any given day at every modelled facility. To enforce HRH constraints on the delivery of these requested health services, the model relies on two key assumptions: A) the standard patient-facing time required from each medical cadre to deliver each type of HSI, and B) The total patient-facing time available from each cadre at each facility on any given day. Based on these assumptions, the model can ensure that HSIs are only delivered until exhaustion of available capabilities, as discussed in detail in [19]. In the model, the former (A) is informed by consensus estimates on the time needed for the delivery of each service as reported by the Human Resources for Health Strategic Plan (HRH SP) 2018–2022 [20], while the latter (B) is informed by the Detailed Annex for the Health Workforce Interventions of the Malawi Health Sector Strategic Plan (HSSP III) for 2023-2030 (HSSP III HRH Annex) [21, 22].

While these sources provide the most reliable theoretical expectation of both A) and B) in the country, a number of factors may alter them in practice (see discussion in [19]). For example, the level of experience of HCWs may be reasonably expected to impact the duration of appointments, while HCWs may cope with a large volume of requests for treatment by shortening the expected appointment duration and/or working overtime. Combined, these and other factors may result in a “productivity level” which differs from the one expected from assumptions around A) and B) alone.

In order to capture these effects while still enforcing HRH constraints in this analysis, we adopt the following approach. The *Thanzi La Onse* model is calibrated to epidemiological and health system data in the period 20152019, and reproduces the observed number of HSIs delivered by the healthcare system during this period [6]. By multiplying the number of HSIs delivered with the assumed time requirement from each medical cadre for each type of HSI (averaged over a year), we can estimate the average number of minutes of patient-facing time *e*ff*ectively* delivered by each HCW type on any given day of that year. This represents the “effective” HRH capabilities available in the country, which indeed surpass those estimated by the HSSP III HRH Annex [22]. Finally, we replace assumptions around B) with the so-estimated effective capabilities in 2018, the year before capabilities expansion is considered, and in doing so account for the real-world productivity of health workers in service delivery (as opposed to theoretical productivity levels expected), while enforcing HRH constraints in subsequent years.

## Appendix B. Effect of consumable availability

In the analysis presented we relied on an assumption of perfect consumable availability over the entire period considered. In this section, we conduct a sensitivity analysis to establish the impact that deviations from this assumption would have on the results. To do so, we evaluate the same scenarios under an assumption of present-day consumable availability (see [6] for details).

In Fig. B.4, we compare how total DALYs incurred scale with yearly expenditure growth under the two assumptions of consumable availability. The return in health as a function of expenditure, which as expected is systematically lower in the case of present-day consumable availability, is largely unaffected by the assumption of consumable availability under the two assumptions. As a result of the higher health burden incurred under a present-day consumable availability, however, the percentage loss incurred under the scenarios forecasted by the IHME is slightly lower than in the case of perfect consumables (12.7 (11.4 – 13.9 at 95% CI) and 5.4 (4.4 – 6.0 at 95% CI) for the lower and upper bound IHME forecasts respectively).

In Fig. B.5, on the other hand, we consider the impact of consumable availability on the evolution of the health burden as a function of the yearly expenditure growth for important areas of health.

In the case of HTM, the effect of consumable availability on the health burden is modest. This is due to the already high consumable availability for diseases mainly funded through vertical programmes [23]. It does however suggest that, should present-day consumable availability levels persist throughout the entire period, a minimum of “< GDP growth” expenditure should be achieved to prevent a reversal of gains in this area.

In the case of RMNCH, the impact of consumable availability is far more evident, such that none of the expenditure scenarios considered would be able to stabilise the health burden in this area if not accompanied by significant invest in consumable access. This is due to generally low levels of consumable availability in this area of health [23].

Finally, in the case of NCDs, the effect of consumable availability is mostly negligible, suggesting again that reach and scope of currently implemented services may be a more significant barrier to an effective translation of expenditure into health outcomes (see discussion in section 3.2).

It may be expected that, in reality, a transition between a present-level and perfect consumable availability would be gradual and a function of the yearly expenditure growth considered. We postpone these considerations to a future analysis.

## Appendix C. IHME forecasts of health expenditure

Two of the scenarios considered in our analysis, namely the “< GDP growth” and “<< GDP growth” scenarios, seek to approximate the lower and upper bounds in the projections of the IHME (see section 2). In this section, we discuss how we obtained these approximations.

The IHME provides projections on the fraction of total GDP allocated to health expenditures in Malawi by four financing source categories: government, DAH, out-of-pocket, and prepaid private, shown in Fig. C.6. In this work, we focus solely on the combined contribution of government and DAH expenditure, and therefore only consider those sources combined. Because our scenarios assume, for simplicity and interpretability, that *g*_fHE_ is constant over the period considered, we average the fractional change in the IHME projections in order to get an equivalent estimate of *g*_fHE_ at the end of the period, in 2040.

Included in the IHME forecasts is emergency COVID-19 funding for the years 2020–2021. The COVID-19 pandemic is however not included in the TLO model, therefore when calculating the average fractional change in *f*_HE_ from the IHME forecasts we extrapolate backwards past 2022 in order to “smooth-out” this emergency funding, as shown by the dashed lines in Fig. C.6. The values obtained from these projections are *g*_fHE_ ∼-1.4 and -3% for the upper and lower bound respectively, which are approximated by our “< GDP growth” and “< GDP growth” scenarios respectively, as shown in the same figure.

**Figure B.4.**
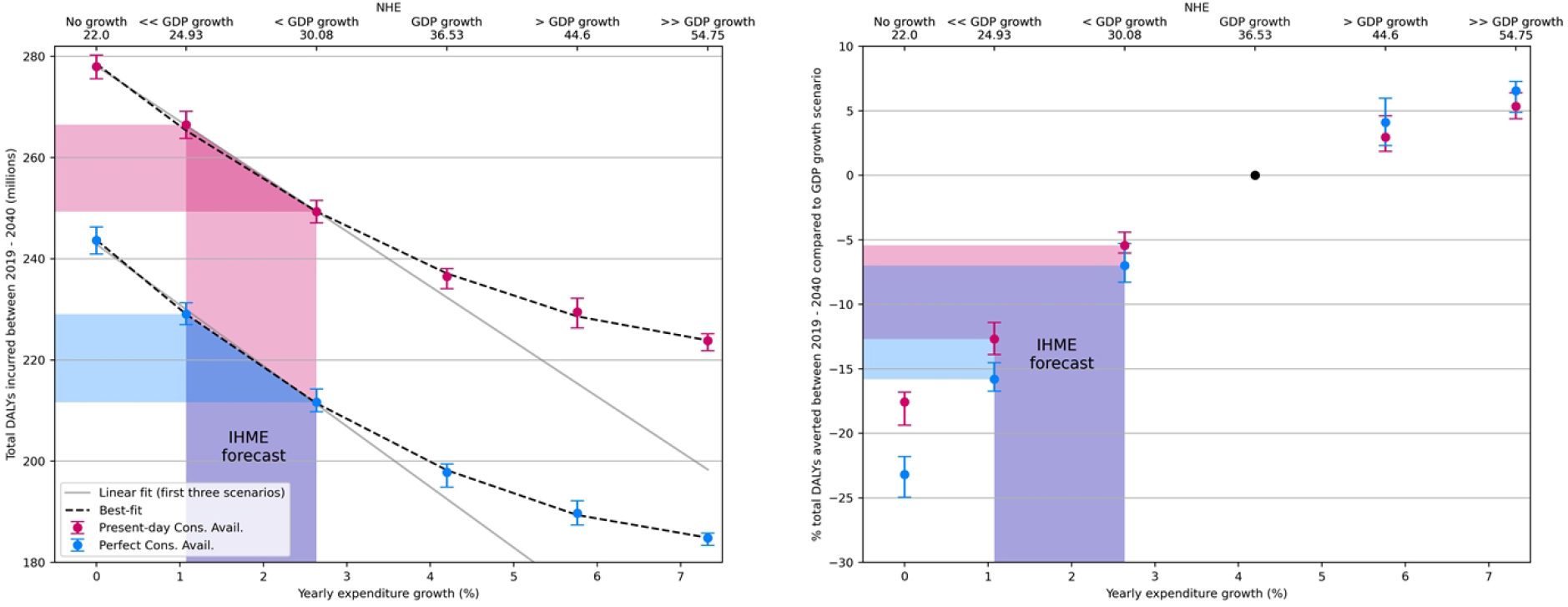
*Left plot:* Total DALYs incurred in the period 2019–2040 (inclusive) as a function of the yearly expenditure growth, as well as the normalised total expenditure (NHE) over that period under each scenario (top x-axis), as defined in Eqn. 2, for two cases of consumable availability: present-day (magenta points) and perfect (blue points) consumable availability. *Right plot:* Percentage DALYs averted in the same period compared to the consumable-availability specific “GDP growth” scenario.

**Figure B.5.**
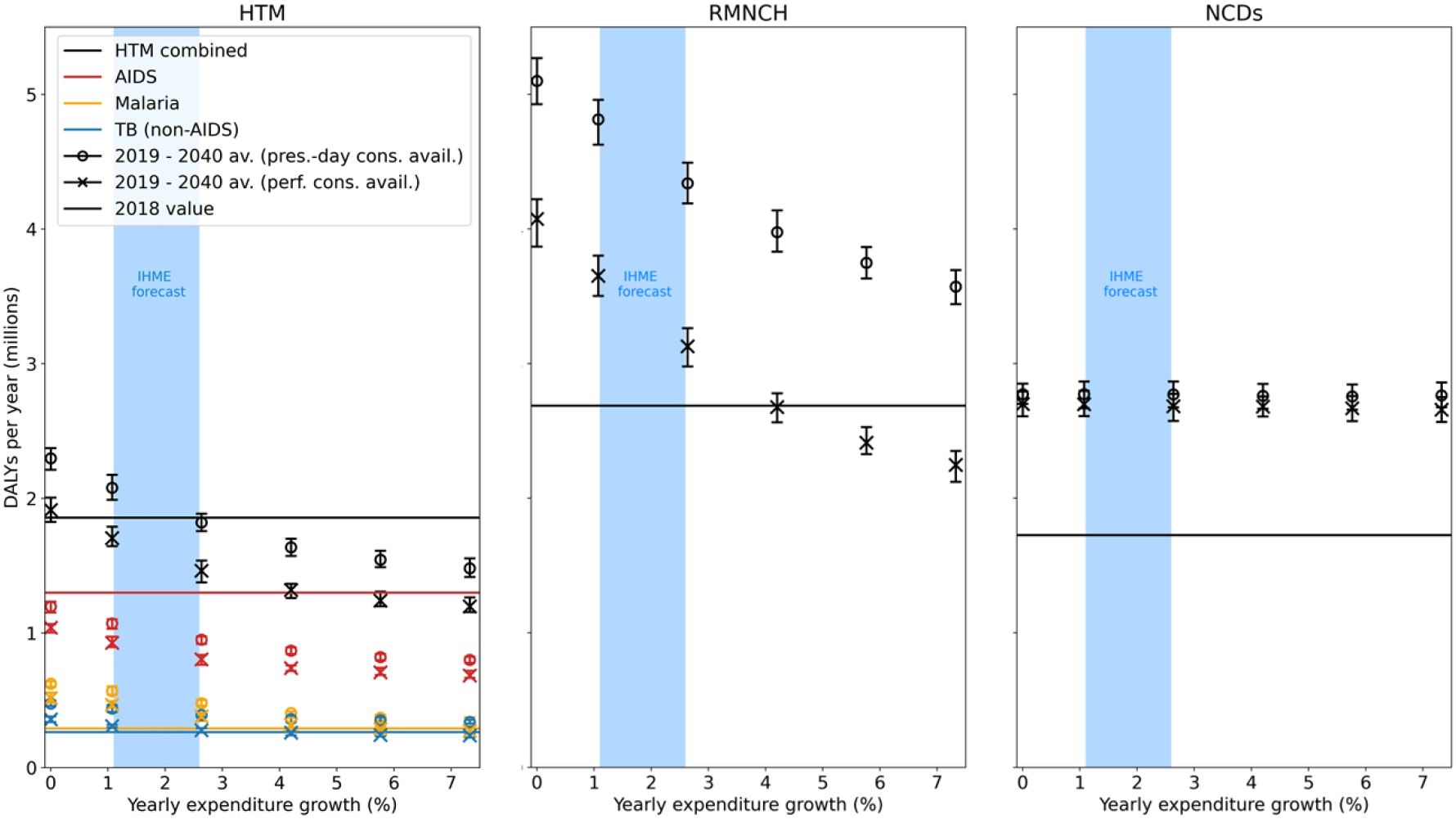
Average yearly DALYs incurred between 2019-2040 (inclusive), grouped into three meaningful categories (see section 3.2), for two cases of assumed consumable availability: the circles show the present-day consumable availability case, while the crosses show the perfect consumable availability case. The straight lines indicate the yearly DALYs burden for each cause in 2018. This means that any scenario *above* the respective 2018 level incurred, on average, a worsening of the health burden due to that cause over the 2019-2040 period, whereas any scenario *below* the respective 2018 level incurred an improvement.

**Figure C.6.**
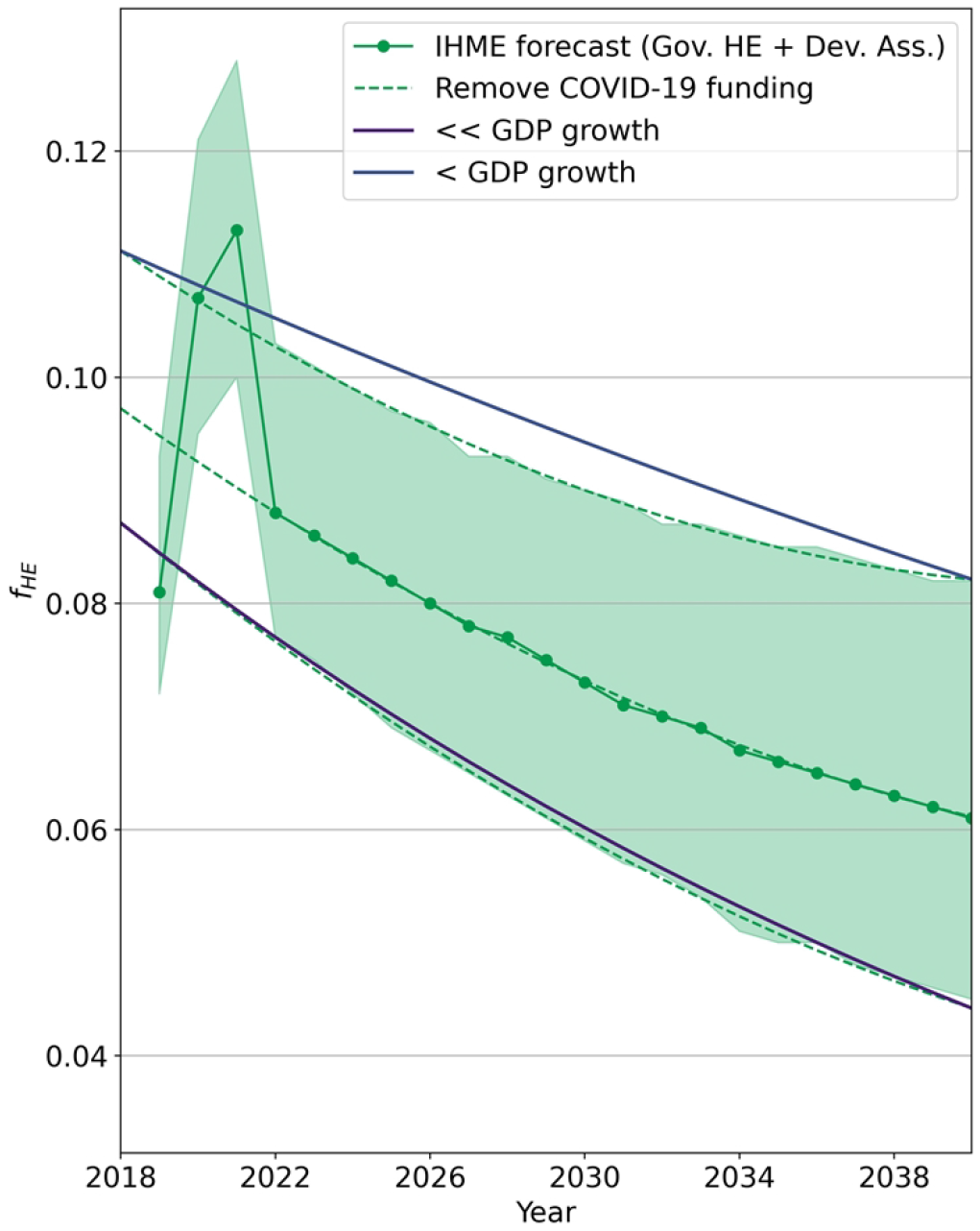
Dashes lines show the best-fit to the mean, upper, and lower bounds of the projections for the combined government and DAH expenditures, extrapolated backward in time between 2018 and 2021 to “smooth out” COVID-19 emergency funding. The fractional change in the upper and lower interpolations are averaged over the entire period to produce the two scenarios that will approximate IHME forecasts in our analysis (“<< GDP growth” and “< GDP growth” respectively).

## Funding

This project is funded by The Wellcome Trust (223120/Z/21/Z) and contributed to the salaries of MM, BS, and TM. MM, BS, TM, and TBH acknowledge funding from the MRC Centre for Global Infectious Disease Analysis (reference MR/X020258/1), funded by the UK Medical Research Council (MRC). This UK funded award is carried out in the frame of the Global Health EDCTP3 Joint Undertaking. The funders had no role in study design, data collection and analysis, decision to publish, or preparation of the manuscript.

## Data Availability

The *Thanzi La Onse* model is open source and available for review and usage at https://github.com/UCL/TLOmodel. In particular, the outputs analysed in this study can be reproduced from model tag “Molaro_et_al_Impact_of_Declining_DAH” (accessible at https://github.com/UCL/TLOmodel/tags).

## Footnotes

[1 All relevant documentation on model assumptions for specific diseases can be found at https://www.tlomodel.org/writeups.html

## Notes

### Competing Interest Statement

The authors have declared no competing interest.

### Funding Statement

Yes

### Author Declarations

The Thanzi La Onse project received ethical approval from College of Medicine Malawi Research Ethics Committee (COMREC, P.10/19/2820) in Malawi. Only publicly available anonymised secondary data is used in the Thanzi La Onse model therefore, individual informed consent was not required.

